# The potential impact of including pre-school aged children in the praziquantel mass-drug administration programmes on the *S.haematobium* infections in Malawi: a modelling study

**DOI:** 10.1101/2020.12.09.20246652

**Authors:** Iwona Hawryluk, Tara Mangal, Andrew Nguluwe, Chikonzero Kambalame, Stanley Banda, Memory Magaleta, Lazarus Juziwelo, Timothy B. Hallett

## Abstract

**Background:** Mass drug administration (MDA) of praziquantel is an intervention used in the treatment and prevention of schistosomiasis. In Malawi, MDA happens annually across high-risk districts and covers around 80% of school aged children and 50% of adults. The current formulation of praziquantel is not approved for use in the preventive chemotherapy for children under 5 years old, known as pre-school aged children (PSAC). However, a new formulation for PSAC will be available by 2022. A comprehensive analysis of the potential additional benefits of including PSAC in the MDA will be critical to guide policy-makers.

**Methods:** We developed a new individual-based stochastic transmission model of *Schistosoma haematobium* for the 6 highest prevalence districts of Malawi. The model was used to evaluate the benefits of including PSAC in the MDA campaigns, with respect to the prevalence of high-intensity infections (> 500 eggs per ml of urine) and reaching the elimination target, meaning the prevalence of high-intensity infections under 5% in all sentinel sites. The impact of different MDA frequencies and coverages is quantified by prevalence of high-intensity infection and number of rounds needed to decrease that prevalence below 1%.

**Results:** Including PSAC in the MDA campaigns can reduce the time needed to achieve the elimination target for *S. haematobium* infections in Malawi by one year. The modelling suggests that in the case of a lower threshold of high-intensity infection, currently set by WHO to 500 eggs per ml of urine, including PSAC in the preventive chemotherapy programmes for 5 years can reduce the number of the high-intensity infection case years for pre-school aged children by up to 9.1 years per 100 children.

**Conclusions:** Regularly treating PSAC in the MDA is likely to lead to overall better health of children as well as a decrease in the severe morbidities caused by persistent schistosomiasis infections and bring forward the date of elimination. Moreover, mass administration of praziquantel to PSAC will decrease the prevalence among the SAC, who are at the most risk of infection.

## Background

Schistosomiasis is one of the Neglected Tropical Diseases, caused by parasitic worms of the genus Schistosoma. All types of schistosome flukes have similar life cycles, requiring freshwater environment and intermediate snail hosts (1,2). People get infected by contact with water containing free-living cercariae. It is estimated that over 230 million people worldwide are at risk of the schistosomiasis infection, contributing to around 100,000 Years Lived with Disability annually (3). Studies have shown that around 90% of all schistosomiasis cases happen in Africa, with sub-Saharan Africa experiencing the highest burden (4).

For the urogenital infections, caused by *S. haematobium*, the most common symptoms are haematuria (blood in urine) and dysuria (painful urination). Prolonged infections lead to a number of complications, including bladder wall pathologies, hydronephrosis and renal dysfunctions, which can progress into bladder cancer or severe kidney failure. The infections may cause infertility in women and increase the risk of acquisition of HIV and HPV viruses (5–7).

As the burden of schistosomiasis is the highest among the school aged children (SAC; 5-15 years old), the burden among the pre-school aged children (PSAC; < 5 years old) is often overlooked (8). Although it is hypothesised that morbidity relates to the worm burden and that light infections, defined as having under 500 eggs per ml of urine, are likely to be asymptomatic in adults, there is little evidence of how the worm burden relates to morbidity in PSAC (9). A few studies have hypothesised that the severe and irreversible complications in adulthood are a consequence of a chronic worm burden during childhood (9). Moreover, schistosomiasis infections, even of relatively low intensity, can cause anaemia, malnutrition and stunted intellectual and physical growth.

Despite the burden of infection in PSAC, they are not included in the mass drug administration (MDA) campaigns as the current formulation of praziquantel has not been approved for use in large-scale preventive chemotherapy of children under 5 years old (10,11). However, a new paediatric praziquantel is currently in clinical trials and is anticipated to be licensed for use by 2022 (12). The release of the paediatric praziquantel raises a question on the potential implications of including PSAC in the MDA campaigns. Mathematical modelling is typically used in such situations to simulate the possible outcomes in various treatment scenarios. For schistosomiasis, such approaches have been used, for example, to advise on the MDA requirements for eliminating the infection worldwide (13), to predict the impact of introducing vaccinations against *S*.*mansoni* infections (14), or to evaluate the cost-effectiveness of the community-wide mass treatment (9).

According to a recent study, which analysed data on schistosomiasis infections in 11 countries, the substantial effort devoted to eliminating schistosomiasis in Malawi resulted in reaching the ‘elimination as a public health problem’ WHO target for the *S*.*mansoni* infections (prevalence of high-intensity infections < 1% in all sentinel sites) and the morbidity control target for *S*.*haematobium* (prevalence of high-intensity infections < 5%) (15). Although schistosomiasis infections affect mostly children, about 80% of employment in Malawi is in the agricultural sector, and therefore a large fraction of the adult population is still at risk of being exposed to the infested water reservoirs (16).

In this study, we analyse the potential impact of including PSAC in the mass drug administration programmes in Malawi by looking at the potential to reach the WHO elimination target in SAC and the change in the prevalence of the high intensity infections in PSAC with MDA coverage and frequency. We build upon an existing multi-disease model framework which simulates lifetime health and health system interactions of the Malawian population. Our focus is on the 6 districts with the highest prevalence of *S*.*haematobium* infections.

## Methods

### Transmission model

The transmission model developed in this project has an individual-based stochastic structure. Each individual has an underlying propensity to become infected, termed the ‘harbouring rate’, drawn from a gamma distribution with scale parameter = 1. The harbouring rate is set this way in order to match the data on the clustering of the worm burden in certain individuals. The shape parameter *k*_*district*_ of the gamma distribution is calibrated to every district and independent of any other individual properties, such as age or place of residence within the district, as described in more detail further in this manuscript. The contribution to the infectious material reservoir and the frequency of infectious contacts with the water is governed by the age-dependent exposure rate *β*_*age*_.

At each time step (1 month), the size of the infectious reservoir is calculated by summing up each individuals’ contributions, that is their current worm burden (WB) multiplied by the exposure rate. The full lifecycle of the schistosomes is simplified and subsumed into the district-dependent parameter *R*_*0*,_ defined here as the average number of female offspring produced by a female worm that effectively infects the definitive human host and survives to maturate into adults. New worms are acquired according to the Poisson distribution, with a rate dependent on the exposure and harbouring rates and the size of the reservoir in the district. The overall distribution of the worm burden in the population, arising from mixing the Poisson and gamma distribution this way is negative binomial, which agrees with the data from the autopsy studies (17). The worms mature to adulthood within 25-30 days and die naturally after 6 years unless they are killed by praziquantel during the mass drug administration before that.

Births and deaths due to any cause are informed by data from United Nations fertility and mortality estimates (18) and Malawi Population and House Census from 2018 (19). We assume no increase in the death rate with schistosomiasis infection, and no immunity gained due to repeated infections. The outputs have a full age structure, but for the presentation of the results, they have been grouped into the 3 age groups: PSAC (0 to 4 years old), SAC (5 to 14 years old) and Adults (15 to 100 years old).

Additional details of the transmission model are available in the Supplementary Information. All parameters used in the transmission model are given in Table 1 and Table 3.

**Table 1.** Parameters for the S. haematobium transmission model.

| Parameter | Parameter description | Value | Reference |
| --- | --- | --- | --- |
| k | Shape parameter of the gamma distribution, used to assign the harbouring rates | Depending on the district | Calibrated, see Table 3 |
| $R_0$ | average number of female offspring produced by a female worm | | |
| $\beta_{PSAC}$ | Age-dependent exposure rate | 0.3 | (20) |
| $\beta_{SAC}$ | | 1 | |
| $\beta_{Adults}$ | | 0.05 | |
| Maturation period | Time from infecting an individual to reach adulthood | 25-30 [days] | (21) |
| Worm lifespan | Adult worm life expectancy | 6 [years] | (22) |

### Calibrating parameters

Parameters *k* and *R*_*0*_ were calibrated to the given prevalence of every district following the methodology from previous studies (22,23). For the calibration, we used the baseline prevalence data obtained from the ESPEN database (24) and filtered for Malawi, years 2002-2012, that is prior to the MDA campaigns being rolled out in the country. We assumed a linear relationship between the parameter k and the baseline prevalence, as reported previously for soil-transmitted helminths (STH) (23), with the slope coefficient fitted using a grid search method. The details of the calibration are provided in the Supplementary Information. Where data for multiple locations within one district were found, an unweighted mean was used in the model as prevalence. The questions we set to answer consider only the six districts with the highest prevalence according to the most recent data available: Blantyre, Chiradzulu, Mulanje, Nkhotakota, Nsanje and Phalombe.

### High-intensity infections

Clinically, a high-intensity infection is recognised as one in which the individual has a measurable number of eggs per ml of urine (epml) that exceeds a certain threshold. The WHO proposes a threshold of 500 epml for *S*.*haematobium* infections (10). Our model tracks the number of worms carried by each individual, not the number of eggs excreted. However, we assume that the number of eggs excreted will be related linearly to the WB and that male and female worms have the same characteristics in all respects, such that we can relate the threshold epml, to a threshold WB, as:

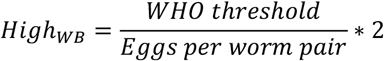

We assume that each worm pair causes a net output of 52 epml (20). Applying the above equation, the high-intensity infection threshold for urogenital infections is equivalent to a WB of 20 worms in our model.

### Mass drug administration

In our model, the individuals selected for treatment are sampled randomly from each age group according to the reported coverage. We assume 100% efficacy of praziquantel in killing adult worms a 100% systematic compliance.

For years 2015 – 2018 the data regarding the coverage of MDA programmes in every district of Malawi, collected and shared by The Department of Schistosomiasis and Soil-Transmitted Helminths (Malawi Ministry of Health, Community Health Science Unit) were used in the model (data is available in the Supporting Information). The data consist of the coverage of the school-based programmes and community treatment. PSAC were not included in the programme, as per the WHO guidelines (10)After 2019, the coverage and frequency of MDA are varied. The details of the MDA events executed in the simulations are listed in Table 2.

**Table 2.** MDA events details. The compliance and efficacy was assumed to be 100% in all scenarios analysed.

| MDA events | Reference | Treatment frequency | PSAC coverage |
| --- | --- | --- | --- |
| MDA in each district in 2015 -2018 (SAC and Adults) | MDA Malawi data; for 2016 the coverage data were interpolated from years 2015, 2017, 2018 Coverage per age group and district is given in the Supplementary Information. | Annual | 0% |
| MDA in each district in years following 2018 | Simulated scenarios | Annual | 0%, 25%, 50% |
|  |  | Biannual | 0%, 50% |

**Table 3.**
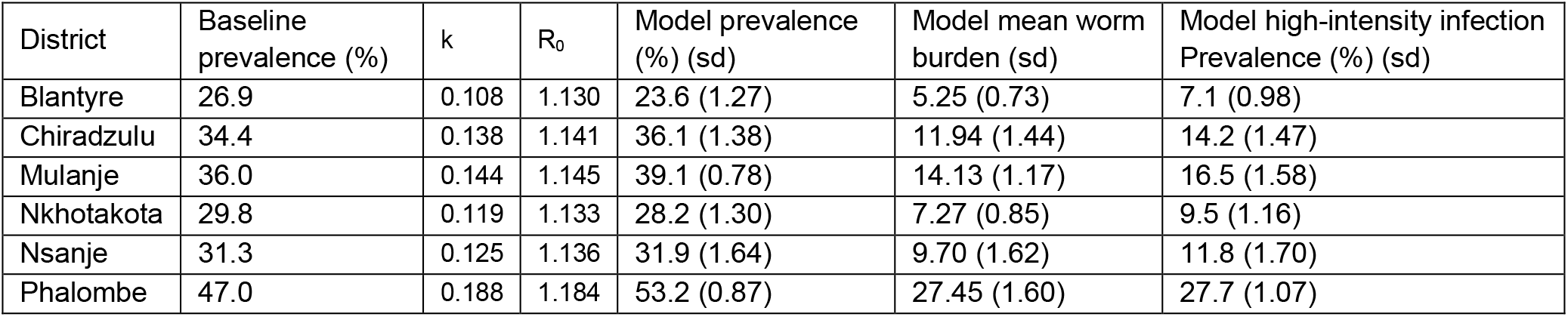
Average prevalence and mean worm burden per district after running 12 simulation for 25 years, without MDA, standard deviation given in brackets. Baseline prevalence was obtained from (24). Parameters k and R_0_ were calibrated to the baseline prevalence as described in the Methods. The last three columns show the outputs of the model: prevalence, mean worm burden and prevalence of high-intensity infections.

| District | Baseline prevalence (%) | $k$ | $R_0$ | Model prevalence (%) (sd) | Model mean worm burden (sd) | Model high-intensity infection Prevalence (%) (sd) |
| --- | --- | --- | --- | --- | --- | --- |
| Blantyre | 26.9 | 0.108 | 1.130 | 23.6 (1.27) | 5.25 (0.73) | 7.1 (0.98) |
| Chiradzulu | 34.4 | 0.138 | 1.141 | 36.1 (1.38) | 11.94 (1.44) | 14.2 (1.47) |
| Mulanje | 36.0 | 0.144 | 1.145 | 39.1 (0.78) | 14.13 (1.17) | 16.5 (1.58) |
| Nkhotakota | 29.8 | 0.119 | 1.133 | 28.2 (1.30) | 7.27 (0.85) | 9.5 (1.16) |
| Nsanje | 31.3 | 0.125 | 1.136 | 31.9 (1.64) | 9.70 (1.62) | 11.8 (1.70) |
| Phalombe | 47.0 | 0.188 | 1.184 | 53.2 (0.87) | 27.45 (1.60) | 27.7 (1.07) |

### Summary of the analysis

The analysis was split into three parts. Firstly, the simulations were run for 25 years simulation time in order to achieve equilibrium and validate the model by comparing the estimated prevalence with the baseline ESPEN data (24). Secondly, to explore the possible timelines of reaching the WHO elimination target, we looked at the prevalence of the high-intensity infections among SAC in each of the MDA scenarios, as specified in Table 2. We then extracted the number of MDA rounds required to decrease the prevalence of high-intensity infections under 1%. Finally, to investigate the impact of the MDA on the high-intensity infections in children, we performed a sensitivity analysis, in which we re-run the simulations with varied WB threshold, above which the infection can be defined as high-intensity infection. The outcomes were quantified by looking at the change in the prevalence of high-intensity infections among PSAC, number of rounds needed to bring that prevalence below 1%, and calculating high-intensity infection years averted in each MDA scenario.

In all simulations, the population size was 10,000 people, split across the 6 districts under analysis (Blantyre, Chiradzulu, Mulanje, Nkhotakota, Nsanje and Phalombe) weighted by the population size and preserving the demographic profile of each district. The results are combined for all 6 districts under analysis. For every MDA scenario, we ran 3 simulations with the same coverage and frequency, as specified in Table 2, and focus on the average outcome.

## Results

### Model validation

The distribution of the worm burden in the population after 25 years of simulation time is shown in Figure 1. The worm burden is negative-binomially distributed, agreeing with autopsy studies (17), with the peak of the mean worm burden (MWB) observed for the SAC (5-15 years old), as expected. The differences between the age groups are much higher in the MWB than prevalence, which is consistent with the previous findings (22). The prevalence in each of the 6 districts produced by the model presents a good fit to the data (Table 4).

**Table 4.** Number of treatment rounds needed to reach the 1% prevalence of high-intensity infections elimination target for SAC in simulations with annual and biannual MDA.

| PSAC coverage | Annual MDA | Biannual MDA |
| --- | --- | --- |
| 0% | 3 | 2 (1 year) |
| 25% | 2 | Not analysed |
| 50% | 2 | 1 (0.5 year) |

**Figure 1.**
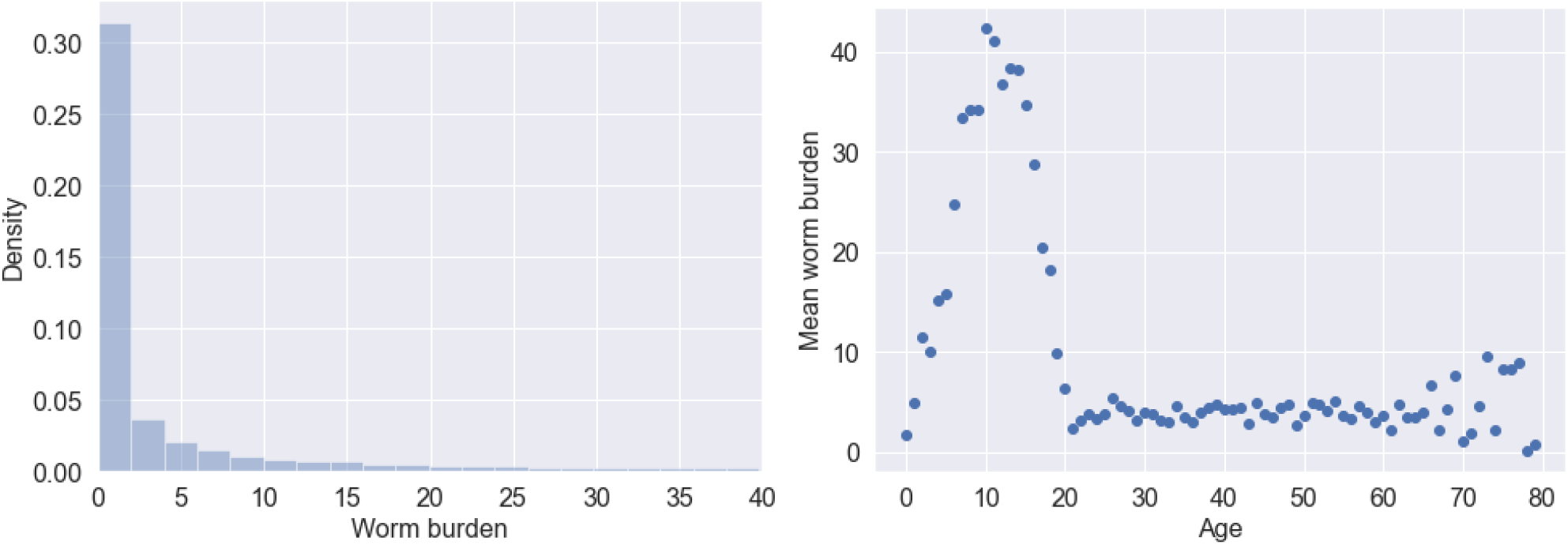
Worm burden generated by the model after running simulations for 25 simulation-years. (Left) Distribution of the worm burden in the total population. The fitted negative binomial distribution has a clumping parameter k = 0.125 and mean = 13.443 (fitting performed with R statistical software); the clumping parameter is comparable with the k-value from other studies (20,22). (Right) Age profile of the mean worm burdens in the entire simulated population.

### The effect of including PSAC in MDA on reaching the elimination target

To examine whether including PSAC in the mass treatment can accelerate reaching the WHO elimination goal, we looked at the predicted prevalence of high-intensity infections in SAC (Figure 2) and the number of treatment rounds needed to achieve the prevalence of high-intensity infections less than 1%, shown in Table 4. If the treatment happens annually, including PSAC with the coverage of just 25% caused the SAC to reach the elimination target within the first 2 additional treatment rounds, as opposed to 3 when PSAC were not included.

**Figure 2.**
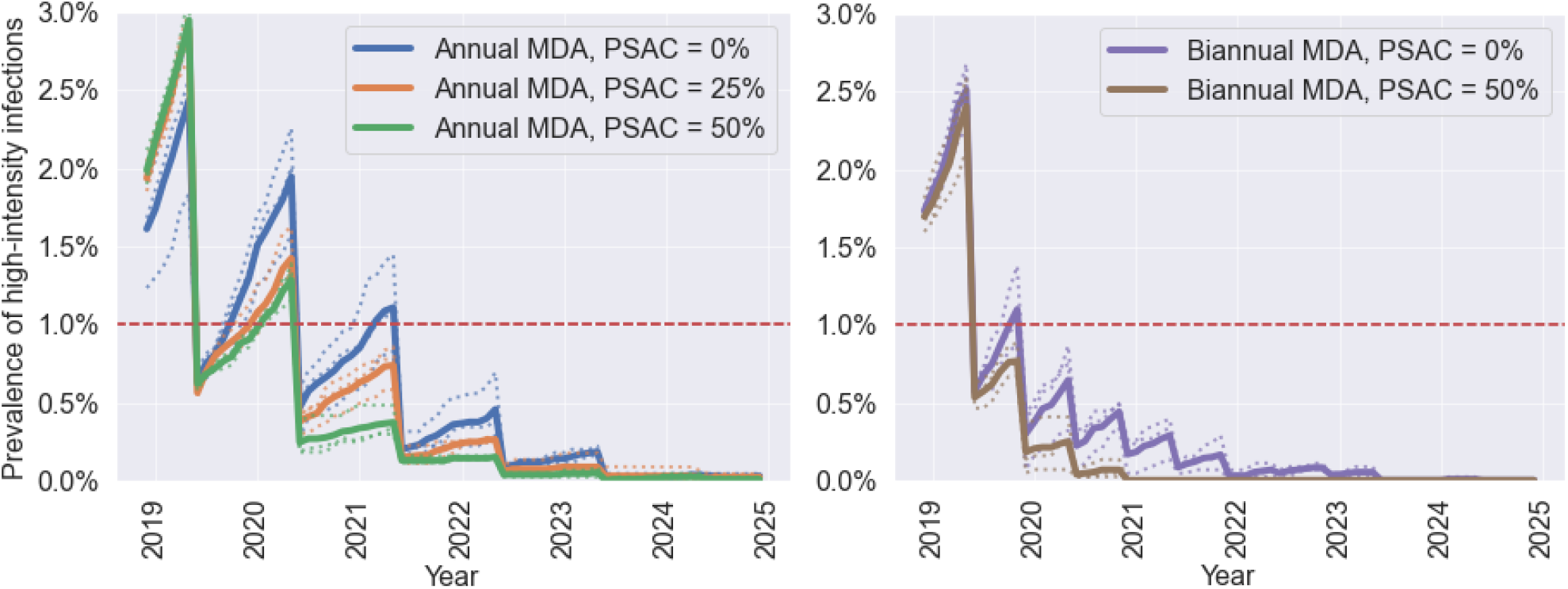
Prevalence of high-intensity infections (over 500 epml, corresponding to around 20+ worms) in SAC depending on the MDA strategy. In all simulations, SAC coverage was fixed to 80% and Adults coverage to 50%. The PSAC coverage was varied between 0% and 50%, as indicated by the plot’s legends. Plot on the left shows simulated outcomes for the annual MDA, and the plot on the right for the biannual MDA. The red line denotes the 1% high-intensity infections prevalence, that is the elimination as a public health problem target defined by the WHO.

**Figure 3.**
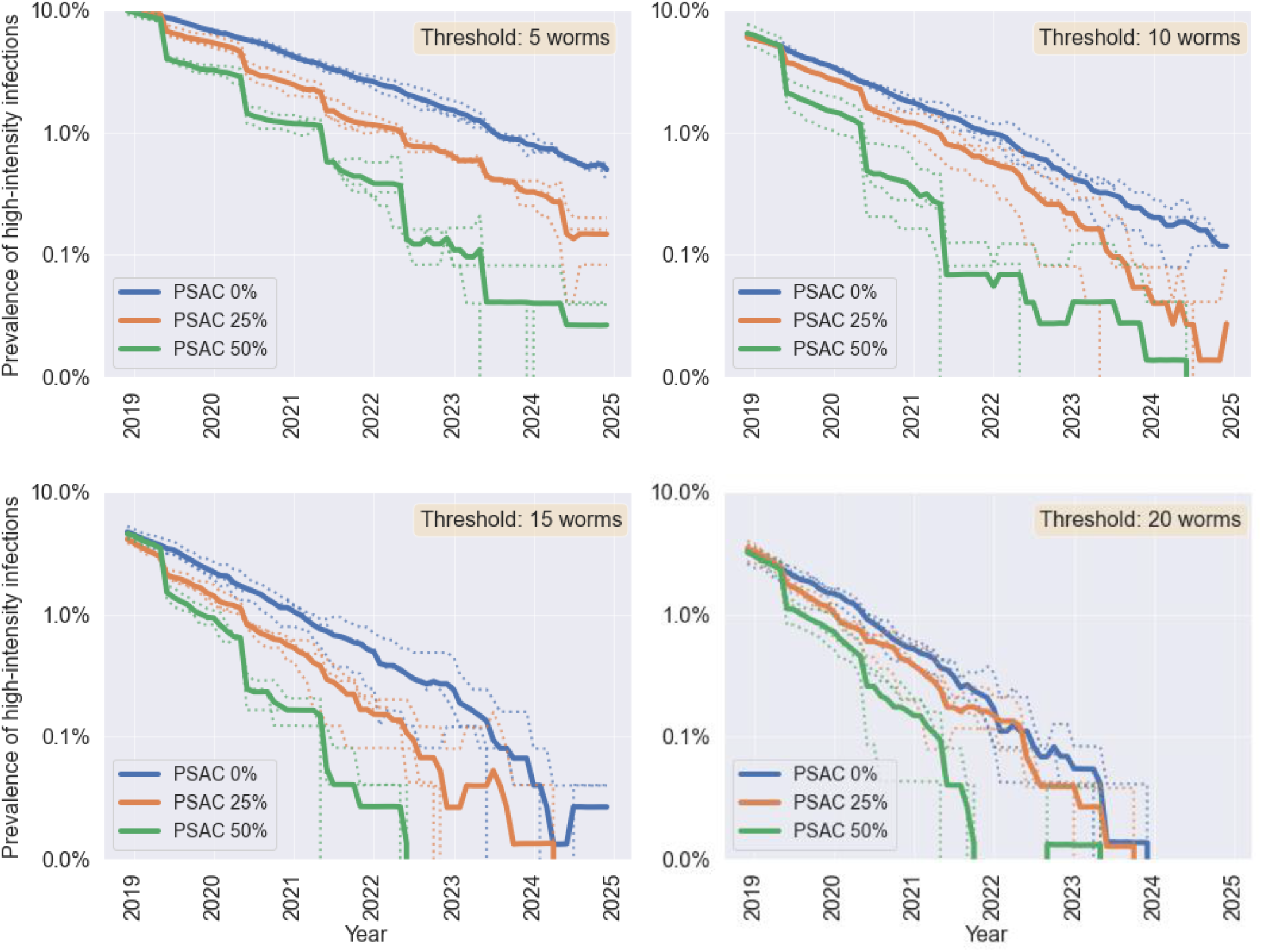
Prevalence of high-intensity infections in PSAC with a worm threshold equal to 5, 10, 15 or 20 worms. Blue line shows scenario of annual MDA with PSAC coverage of 0%, magenta line - coverage 25%, yellow line – coverage 50%.

### High-intensity infections in PSAC

Based on these results, in Table 5 we show the expected number of MDA rounds required to decrease the prevalence of high-intensity infections below 1%. If the current strategy of MDA continues, our model predicts that 5 more rounds will be required to reach 1% prevalence of high-intensity infections among PSAC with the threshold of 5 worms, compared to 2 years if the threshold is the same as for SAC, that is 20 worms. By including the PSAC with the coverage of 25 or 50%, the 1% prevalence target would be reached within 4 or 3 annual treatment rounds respectively in the ‘worst-case’ scenario, that is when the threshold of high-intensity infections among PSAC is 5 worms.

**Table 5.** Rounds of annual MDA needed to decrease the prevalence of high-intensity infections among PSAC below 1%, depending on the threshold of the high-intensity infection.

| Number of worms / PSAC coverage | PSAC 0% | PSAC 25% | PSAC 50% |
| --- | --- | --- | --- |
| 5 worms | 5 | 4 | 3 |
| 10 worms | 3 | 2 | 2 |
| 15 worms | 2 | 2 | 1 |
| 20 worms | 2 | 1 | 1 |

In Figure 4, we show the number of high-intensity infection case years averted, depending on the PSAC coverage in the MDA and the high-intensity infection threshold among PSAC. The case years averted for different MDA strategies vary the most for the lowest threshold of 5 worms. By including PSAC in the annual MDA with 25% coverage, about 5 years per 100 people are averted in 5 years of treatment. This value doubles if the coverage is increased to 50%.

**Figure 4.**
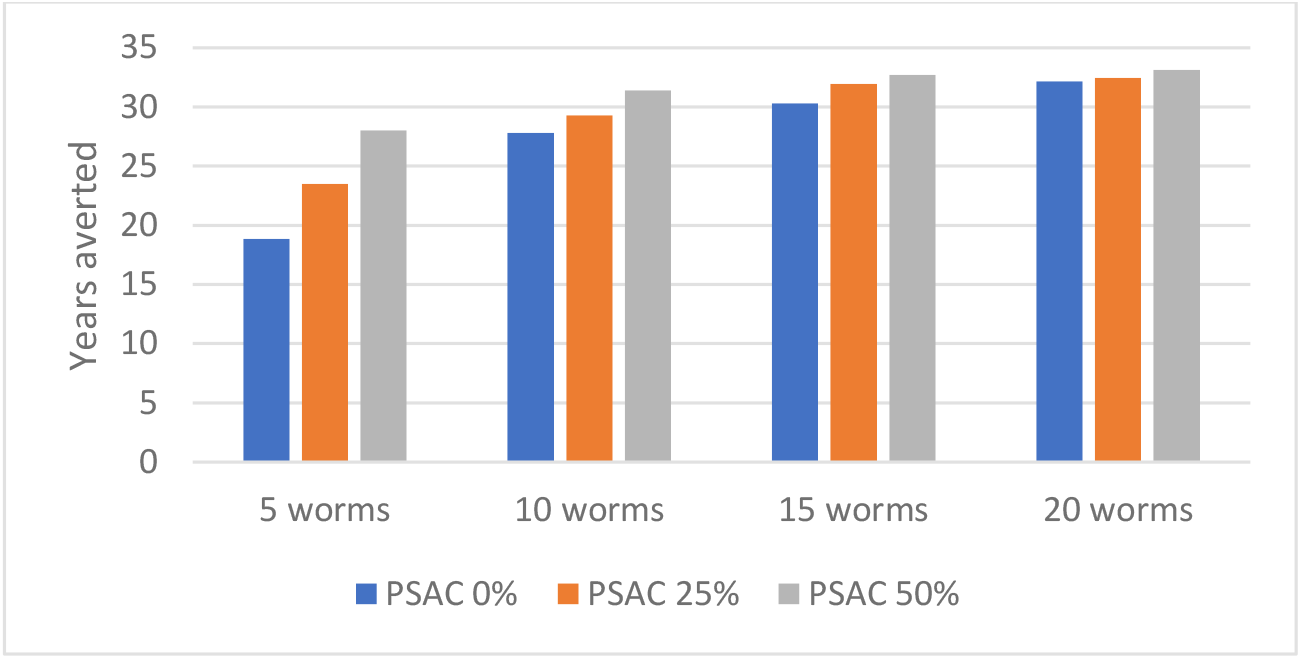
High-intensity infection years averted among PSAC in 5 years of MDA, per 100 children.

## Discussion

We showed that expanding the coverage of MDA to include PSAC can lead to a accelerated decrease in the high-intensity infections in SAC compared to the current strategy. By increasing the PSAC coverage to 25 or 50% or increasing the frequency of MDA to twice per year without including PSAC, the WHO goal of elimination of urogenital schistosomiasis in the highest burden districts of Malawi can be achieved within only 2 additional rounds of MDA.

The sensitivity analysis, focusing on the high-intensity infections among PSAC, indicated the importance of treating the PSAC in mass drug administration, especially if we consider the ‘worst-case’ scenario of 5-worms threshold of high-intensity infection in PSAC. Looking at the high-intensity infection case years averted (Figure 4), the impact of including PSAC in the annual MDA programmes is greatest for the worst-case scenario of the high-intensity threshold of 5-worms. Up to 28 high-intensity infection years per 100 children can be averted within 5 years if the coverage of PSAC is increased, in comparison to 18 years if the current strategy is continued.

Including PSAC in the mass drug administration can have a major impact on controlling morbidity due to urogenital infections, not only among PSAC but other age groups as well. One of the biggest risks of chronic infections in PSAC is that they might lead to severe and permanent symptoms later in life. For example, prolonged schistosomiasis infections may lead to developing serious conditions such as infertility, bladder cancer, kidney failure or inflammation of internal organs (1,5,25,26). Moreover, studies have suggested that prenatal and infant exposure to schistosomiasis and other STH might be correlated to a reduction in the efficacy of childhood disease vaccinations (8). Schistosomiasis infections in childhood have also been found to impair growth and reduce learning abilities, causing long-term and irreversible implications in terms of quality of life in adulthood (1,27–30). Taking all this into account, the benefits of treating the pre-school aged children, while difficult to measure, could be immense.

Although children under 5 years old can be administered praziquantel upon the positive diagnosis of the infection by the laboratory test, studies suggest this does not happen regularly (27). The probable reason for that is the poor understanding of the symptoms of schistosomiasis in young children, which are often non-specific (malnutrition, anaemia, microhaematuria) and easy to mistake for other diseases of poverty or for malaria. This makes the infections extremely difficult to recognise in the early years of the child’s life. Moreover, Bustinduy et al. suggest that the common preconception is that the younger children will be treated with praziquantel later in their life, during a school-based MDA campaign, by which time it might already be too late to reverse the damage (27). Studies conducted in Malawi revealed that schistosomiasis is a common problem in PSAC in rural areas (31,32). Poole et al. examined mothers and their pre-school aged children in 12 villages in the Chikwawa district and showed that around 25% of children had haematuria or albuminuria. The children had regular and extensive contact with the infested water, as they often accompanied mothers in the water during daily activities such as washing clothes (31).

The Global Burden of Disease study from 2017 (33) revealed that dietary iron deficiency was the number one cause for most disability in Malawi, having overtaken HIV/AIDS and increased by 34.5% since 2016. Additionally, in 2017 malnutrition was the prevailing risk factor driving the most death and disability combined. As urogenital schistosomiasis is contributing to both malnutrition and iron deficiency, the paediatric formulation of praziquantel and preventive chemotherapy for the pre-school aged children has the potential of decreasing those two risk factors and consequently reducing the disability levels in Malawi.

Although the control and elimination targets defined by WHO refer only to the high-intensity infections, two things are worth noting. Firstly, the infections are likely to be more pathogenic in the PSAC and infants (9), therefore the same threshold of high-intensity should be used with caution. Secondly, light infections should not be overlooked: their implications are not well understood, and even with a small worm burden the eggs can cause inflammation of the internal tissues (34,35).

The full life cycle of schistosomiasis infection is complex and requires modelling the dynamics of the schistosomes’ various life stages, including the dynamics of the snail populations. In many existing models of schistosomiasis infection, including the model presented here, simplifications are introduced, based on the fact that the lifespans of the adult worms are orders of magnitude longer than the preceding life stages (36). In the model described here, we adopted the main assumptions from the Anderson et al. schistosomiasis model (13) and its stochastic version (37), however certain limitations have to be addressed.

Firstly, the model does not explicitly include the probability of adult schistosomes mating, which is important in the low-prevalence districts. Probability of mating is one of the vital issues that drive the population of schistosomes extinct in the areas with low schistosome densities. Because we disregard this characteristic, our model may be overestimating the worms’ density in the low-burden districts, however in this study we focused on the high-risk districts, in which the probability of mating is relatively high.

Some people have elevated risks of acquiring schistosomiasis infection. This is caused by a variety of biological, environmental and behavioural factors, such as profession or personal hygiene (38,39). In our models, we allocate those risks and predispositions randomly, which might lead to the underestimation of the worm burden in groups with elevated risks. We still include the tendency of children to spend more time in contact with the water by using age-dependent exposure rates to estimate the risk of acquiring new infection, as well as their increased input of the infectious material to the environment. The proximity to the infested water reservoir is also covered to some extent through using observational data from Malawi to estimate the initial prevalence in each of the districts separately and further assuming no mixing of individuals between the districts. Because of this, we preserve the higher risks of infections in high-prevalence districts, which might be caused by a larger density of water reservoirs, rural areas and poverty (8,40).

To estimate the baseline prevalence, we calculated the average prevalence of *S*.*haematobium* infections within each district of Malawi using data often collected from a single school or village. However, schistosomiasis is a highly localised disease dependent on the proximity of the water reservoir with infected snails. As a consequence, prevalence within a district can be highly heterogeneous (25,41,42).

## Conclusions

Our analysis suggests that including pre-school aged children (PSAC) in the MDA campaigns can reduce the time needed to achieve the WHO elimination target in the highest-prevalence districts in Malawi, reaching it one year sooner than with the current strategy. Furthermore, we showed that including the PSAC in the preventive chemotherapy programmes can significantly reduce the number of the high-intensity infection case years for PSAC if their threshold is lower than currently set by WHO guidelines. Although difficult to know with the current knowledge, this might lead to a substantial decrease in the severe morbidities in adult life, such as bladder cancer, infertility or kidney failure, as well as better overall health and development of the children living in the endemic areas. The uncertainties around the high-intensity infections among PSAC highlight the need for more research in the area of the schistosomiasis morbidities and their complications in PSAC.

## Data Availability

All data generated or analysed during this study are included in this published article or the referenced materials. The MDA coverage per year and age group as used in the model is included in the Supplementary Information.

## Declarations

### Ethics approval and consent to participate

Not applicable.

### Consent for publication

Not applicable.

## Competing interests

The authors declare that they have no competing interests.

## Funding

IH, TM and TBH acknowledge funding from the MRC Centre for Global Infectious Disease Analysis (reference MR/R015600/1), jointly funded by the UK Medical Research Council (MRC) and the UK Foreign, Commonwealth & Development Office (FCDO), under the MRC/FCDO Concordat agreement and is also part of the EDCTP2 programme supported by the European Union. TM and TBH were also funded by the Thanzi la Onse grant (RCUK MR/P028004/1).

## Authors’ contributions

Model design and manuscript writing: IH, TM, TBH. Model implementation and simulations: IH. Study design and critical review of the manuscript: all authors.

## Acknowledgments

The authors would like to thank other people involved in the design and implementation of the Thanzi la Onse epidemiological model, including Prof. Andrew Phillips, Dr Timothy Colbourn and Dr Asif Tamuri. The authors would also like to thank Dr James Truscott, Dr Klodeta Kura and Dr Robert Hardwick for advice on the transmission model and Dr Hugo Turner for the advice on the current schistosomiasis treatment strategies worldwide.

## Abbreviations

MDA: mass drug administration
PSAC: pre-school aged children
SAC: school aged children
STH: soil-transmitted helminths
TLO: Thanzi la Onse
WB: worm burden
WHO: World Health Organisation

## Supplementary Information

### Additional file 1

#### Assignment of the new infections

The transmission model is adapted from earlier work detailed in (20,21,23,37).

The main idea behind the schistosomiasis model developed for this project is that every individual has a ‘worm burden’ (WB) which is tracked, and an underlying propensity to become infected, termed the ‘harbouring rate’. At birth, individuals have a zero WB and a harbouring rate is drawn from a gamma distribution.

At any particular time, all individuals with a positive worm burden (WB) contribute to the total worm reservoir, from which people acquire new worms with a rate dependent on their age and harbouring rate. The full lifecycle of the schistosomes is simplified and subsumed into the district-dependent parameter R0. Persons can be infected multiple times, which leads to increases in their WB. High-intensity infections are classed as those in persons where the WB exceeds a defined threshold. The worms can be killed with praziquantel, upon administration of which the worm burden decreases.

New infections are assigned independently in every district. Assignment of new infections happens in steps described below.

1. Calculating the size of the reservoir Every person contributes to the total reservoir of infectious material. We do not model the reservoir explicitly by the number of schistosome eggs, instead the reservoir will indirectly model the force of infection. At each moment, the reservoir is modelled on a district level, by summing all individuals’ contribution to the total worm reservoir, that is the worm burden per person multiplied by the age-dependent exposure rate. The exposure rate allows to accommodate for the increased input of the eggs to the environment by the people who spend more time in contact with water, and who are at the same time more exposed (the value of exposure rate depends only on the age of the individual). This value is then multiplied by R_0_. This is described by the following equation:

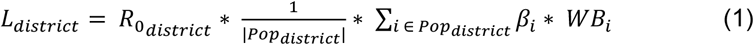 Where: L_district_ = measure of the number of worms per district combined with the average number of infections arising from each worm, R_0 district_ = Basic reproduction number in the district, Pop_district_ = the whole population of individuals in the district, β_i_ = exposure rate of individual *i*, WB_i_ = worm burden of individual *i*. *(suppressing time dimension from the subscripts)*
2. Generating numbers of newly harboured worms For each individual, a number of newly harboured worms is randomly sampled from a Poisson distribution, accounting for the mean worm burden in the district, the harbouring rate of the individual, and their exposure rate, as per the equation below:

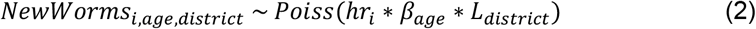 Where: NewWorms_i, age, district_ = Number of new worms acquired by individual *i* of age group *age* and living in district *district*, Poiss() = random variable drawn from a Poisson distribution, hr_i_ = harbouring rate of individual i, β_age_ = exposure rate of age group *age*, L_district_ = mean worm burden in *district*, as per equation (1).
3. Increasing the worm burden We say that a new worm has been successfully harboured if the cercariae managed to penetrate the skin, mature into an adult worm and establish itself in the human host organism. This is a density-dependent process, which can be expressed in terms of a probability of successful establishment of the new worms in the host carrying n-worms, as given in the equation below:

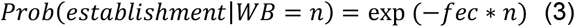 where *fec* is a constant fecundity parameter of the worms, set to 0.005 (20). For every person who is assigned a positive number of new worms, a random variable is generated from *Bernoulli(p)* with probability *p* as given in equation (3). The value of this variable, 0 or 1, determines whether the establishment of the new worms succeeded, and hence the worm burden increases, or failed, in which case the worm burden does not change. The increase in the worm burden for an individual *i* of age *age* with current WB = *n* is effectively drawn from *Poiss*(*hr*_*i*_ * *β*_*age*_ * *L*_*district*_)* *Bernoulli*(−*fec* * *n*).
4. For people whose worm burden increases due to the successful establishment of new worms, the increment happens only after a period of the worms’ maturation, which is taken to be a period lasting 25-30 days (21).

All parameters are fixed throughout the simulation. Their values are given in **Error! Reference source not found**. and 2 in the main manuscript text.

### Additional file 2

#### Calibrating the parameters

Parameters k and R_0_ were calibrated to the given prevalence of every district, as per the equations below, following the methodology from (21,23).

Parameters k (clumping parameter), P (prevalence), MWB and R_0_ are all district dependent, but for clarity, we drop the district-index here. Parameter γ was equal to 0.005 (20).

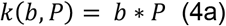

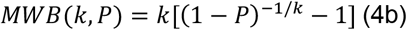

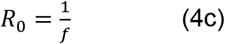

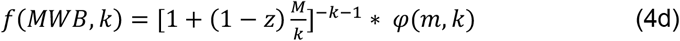

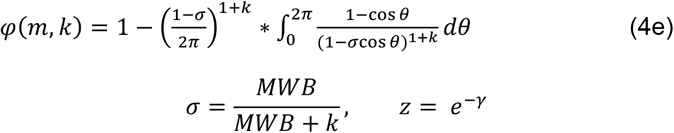

For the soil-transmitted helminths (STH), parameter b in the equation 4a is estimated to be approximately 0.5 (23). For schistosomiasis, this parameter should be less than for the STH, because of the differences in the reproduction processes between those groups of parasites. To find an appropriate value for the parameter b, we performed grid search in the space b ϵ [0.2, 0.3, 0.4, 0.5]. Parameter value b = 0.4 was chosen as a parameter that minimised the difference between the observed data and model outputs.

Applying equations 4a – 4e to the baseline prevalence, we obtain the calibrated parameters k and R_0_ for every district of interest. The results of these calculations are shown in Table 3 in the manuscript text.

### Additional file 3

#### Mass-drug administration coverage data

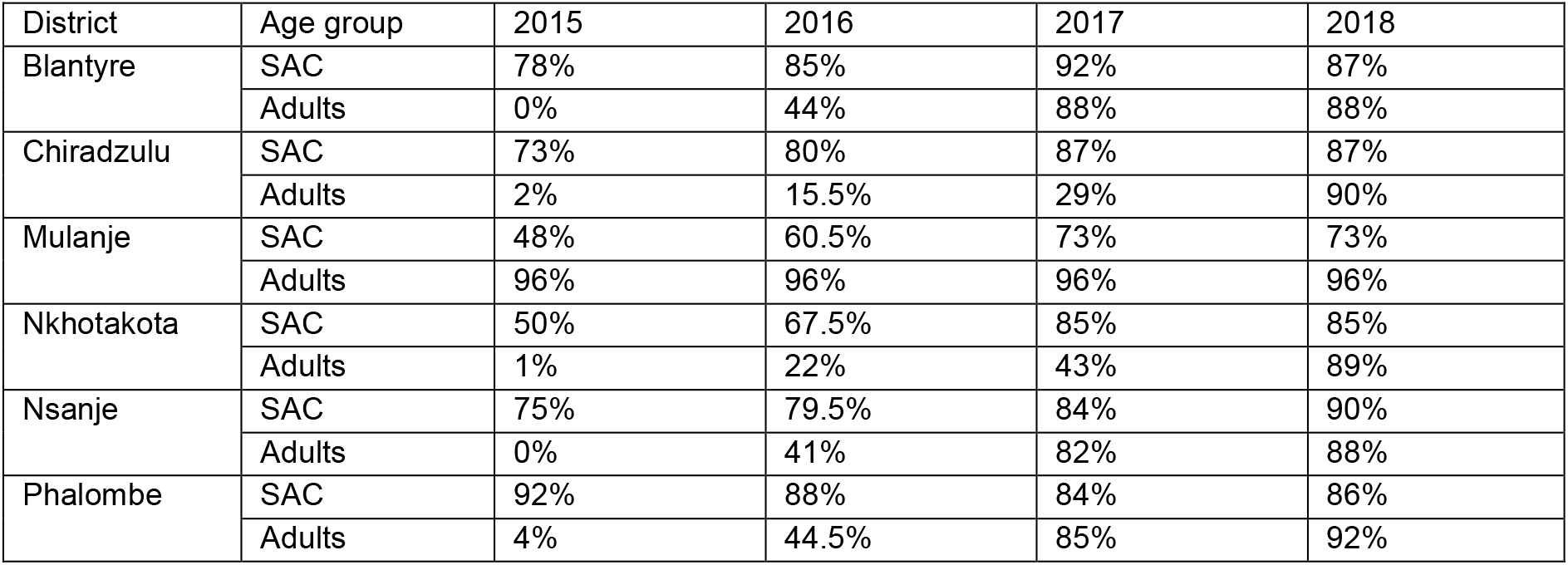

## Notes

### Competing Interest Statement

The authors have declared no competing interest.

### Clinical Trial

This was not a clinical trial

